# Identifying a Heterogeneous Effect of Atrial Fibrillation Screening in Older Adults: A Secondary Analysis of the VITAL-AF Trial

**DOI:** 10.1101/2024.05.17.24307559

**Authors:** Sachin J Shah, Jay M. Iyer, Leila Agha, Yuchiao Chang, Jeffrey M. Ashburner, Steven J. Atlas, David D. McManus, Patrick T. Ellinor, Steven A. Lubitz, Daniel E. Singer

**Author notes:** <u>Contact</u>: Sachin J Shah, MD, MPH, Massachusetts General Hospital, 100 Cambridge St, Suite 1600, Boston, MA 02114.

## Abstract

**Background:** One-time atrial fibrillation (AF) screening trials have produced mixed results; we sought a subset for whom screening is effective.

**Methods:** We conducted a secondary analysis of VITAL-AF, a randomized trial of one-time, brief, single-lead ECG screening during primary care visits. We tested two approaches to identify a subgroup where screening is effective. First, we developed an effect-based model using a T-learner. Specifically, we separately predicted the likelihood of AF diagnosis under screening and usual care conditions; the difference in probabilities was the predicted screening effectiveness. Second, we used a validated AF risk model to test for heterogeneous screening effectiveness.

**Results:** In the effect-based analysis, in the highest quartile of predicted screening effectiveness, AF diagnosis rates were higher in the screening group (4.00 vs. 2.88 per 100 person-years, rate difference 1.12, 95% CI 0.11 to 2.13). In the risk-based analysis, in the highest quartile of baseline AF risk, AF diagnosis rates were also higher in the screening group (5.55 vs. 4.23 per 100 person-years, rate difference 1.32, 95% CI 0.14 to 2.50). Predicted screening effectiveness and predicted baseline AF risk were weakly correlated (Spearman correlation coefficient 0.23). Patients with low primary care use, using rate control medications, females, and Black patients were overrepresented in the high-effectiveness group even when they were not at high risk of developing AF.

**Conclusions:** In a secondary analysis of VITAL-AF, we identified subgroups where one-time screening was associated with increased AF diagnoses using both effect-based and risk-based approaches. In this study, predicted AF risk was only a partial proxy for predicted screening effectiveness. Even when individuals are not in the high-risk subset, features like low primary care use and rate control medication use can identify individuals for whom AF screening has a large impact. Future AF screening efforts should focus on screening both “high-risk” and “high-effectiveness” individuals.

**What is Known:**

- Because trials testing office-based screening for atrial fibrillation have produced mixed results, some have suggested we focus screening efforts on high-risk individuals.
- Newer methods allow us to test for screening heterogeneity using risk-based analyses and separately effect-based analyses, which disentangle screening effects from baseline disease risk.

**What the Study Adds:**

- Both the risk-based analysis and the effect-based analysis identified “high-risk” and “high-effectiveness” subgroups, respectively, where one-time AF screening was effective.
- “High-risk” and “high-effectiveness” groups only partially overlap; even when individuals are not in the high-risk subset, features like low primary care use and rate control medication use an identify individuals for whom AF screening has a large impact.
- Future AF screening efforts should focus on screening both “high-risk” and “high-effectiveness” people.

## INTRODUCTION

The impetus to screen for atrial fibrillation (AF) is clear—AF is common and increases the risk of disabling strokes.^1,2^ Among those 65 years old, the lifetime incidence of atrial fibrillation is 33%.^3^ The goal of screening is to identify cases earlier than usual so stroke-preventive therapies, notably anticoagulation, can be used in appropriate patients. However, randomized clinical trials (RCTs) of screening interventions have produced mixed results, and in 2022, the United States Preventive Services Task Force concluded that there was insufficient evidence to recommend routine screening for AF.^4–8^

One-time screening during routine clinical care is appealing because it is practical and is thought to identify individuals with high-burden AF.^9–11^ Nevertheless, all but one RCT testing one-time screening in traditional care settings (e.g., primary care offices) have failed to show that screening identifies more cases of AF in 6 to 12 months.^5–8^ However, it remains unclear if there is a subset of people for whom screening is effective. While trials typically test for treatment effect heterogeneity one subgroup at a time, newer methods allow for the examination of heterogeneity across multiple factors.^12–16^ Contemporary approaches include risk-based analyses and effect-based analyses.^17,18^ These approaches have been successfully applied in clinical decision models such as the Dual Antiplatelet Therapy (DAPT) score, which guides the duration of dual antiplatelet therapy after coronary stenting, and the PFO-Associated Stroke Causal Likelihood (PASCAL) score, which guides patient selection for PFO closure.^19,20^

Effect-based approaches are compelling because they disentangle treatment effects from baseline disease risk.^13^ In the case of AF screening, screening effectiveness is high when baseline disease risk is sufficiently high, and detection is unlikely through usual care (e.g., due to limited access to care or asymptomatic disease). Screening might be less effective for those at high baseline risk if they also have a high probability of being identified through routine clinical care. Identifying a subset of patients where screening is effective could support future screening strategies, including targeted screening trials.

Our goal was to determine if one-time AF screening during routine clinical care is effective in a subset of older adults. To accomplish this, we conducted a secondary analysis of the VITAL-AF RCT, which demonstrated that single-lead ECG screening for adults 65 years and older during their primary care office visits did not significantly change the rate of new AF diagnoses.^21^ In this secondary analysis, we aimed to identify a subset of people in whom screening is effective using “effect-based” and “risk-based” approaches.

## METHODS

### Study Design and Participants

The data that support the findings of this study are available from the corresponding author upon reasonable request. This is a secondary, post-hoc analysis of the VITAL-AF trial to identify if AF screening is effective in a subset of individuals. The design and primary results from VITAL-AF have been published.^21,22^ In brief, VITAL-AF was a pragmatic, cluster-randomized trial that tested the effectiveness of single-lead ECG screening during primary care visits compared to usual care. The trial randomized 16 primary care practices (8 to screening and 8 to usual care) in the Massachusetts General Hospital Primary Care-Based Research Network between July 2018 and October 2019. We did not adjust for the cluster-randomized design in this study because the intracluster correlation was low (0.0013). For these analyses, individuals 65 years and older without a prior diagnosis of AF presenting for a primary care appointment were included, and participants with missing predictors were excluded (**Figure S1**). All analyses were conducted on an intention-to-screen basis (i.e., group assignment was based on the patient’s first visit to a study practice during the study period). The Mass General Brigham Institutional Review Board approved the research protocol. Participants provided informed consent to participate. The study was considered minimal risk, and a waiver of documentation of informed consent was granted.

### Procedure

As previously described, screening was conducted when consenting patients placed their fingers on a single-lead AliveCor KardiaMobile 30-second ECG device (AliveCor Inc, Mountain View, CA). The screening resulted in one of five possible results, “Possible AF,” “Normal,” “Unclassified,” “No analysis (Unreadable),” and “Patient Declined Screening.” All subsequent clinical management was determined by primary care clinicians, including follow-up 12-lead ECGs. Independent cardiologists reviewed all AliveCor tracings within 7 days and notified primary care clinicians if a prespecified actionable rhythm was identified.

### Outcome

The primary outcome was an adjudicated incident AF diagnosis. Each clinical practice was enrolled for 12 months; participants were followed until the primary care practice to which they belonged completed participation. Thus, follow-up time was measured from each participant’s first visit date until the date their primary care practice completed participation or death, whichever came first. Potentially new AF diagnoses were identified from the electronic medical record using the same approach in both study arms by a centralized data repository.^23^ Specifically, individuals with an International Classification of Diseases, 10^th^ Revision code for atrial fibrillation or flutter or a 12-lead ECG with atrial fibrillation or flutter in the diagnostic statement were identified. These potential new AF diagnoses were then adjudicated by 2 research nurses with a cardiologist unaffiliated with the study resolving differences. The committee adjudicated events as “incident,” “prevalent,” or “not AF.”^24^

### Predictors

We obtained patient characteristics from the electronic medical record, including demographics, medical diagnoses, medication use, physiological measures, and prior health use. We used measures obtained on the participant’s first visit date or the value most immediately prior. Candidate predictors were chosen because of their association with AF, cardiovascular disease, or health care utilization. Demographics included age, sex, and English language preference. Medical diagnoses included hypertension, myocardial infarction, coronary artery disease, diabetes, congestive heart failure, prior stroke, vascular disease, anemia, bleeding history, chronic kidney disease, and tobacco smoking. Medications were grouped into one of the following categories: oral anticoagulants, rate control medications, antihypertensives, or antiarrhythmic medications. Rate control medications included beta blockers, calcium channel blockers, and digoxin; they partially overlapped with antihypertensives. Physiological measures include systolic blood pressure, diastolic blood pressure, heart rate, height, and weight. Healthcare utilization measures included 12 lead ECGs in the prior year and the number of primary care visits in the prior year.

### Effect-based approach to heterogeneity

Our objective was to identify individuals for whom screening is effective by measuring the heterogeneity of AF screening using an effect-based approach.^13^ To do so, we estimated the screening effectiveness for each individual given their observed characteristics, i.e., the conditional average screening effectiveness.^17^ Specifically, we used a T-learner wherein, for each individual, we estimated the probability of the outcome (i.e., new AF diagnosis) had they been randomized to screening and, separately, had they been randomized to usual care.^18^ Because we are conducting a secondary analysis of a pragmatic trial, we describe the output as screening effectiveness. This analysis was operationalized in 3 steps. The process is visualized in **Figure S2**.

#### Step 1: Predict counterfactual AF diagnosis had participants been randomized to the opposite arm

For participants randomized to the screening group, we predicted the probability of AF diagnosis had they been randomized to usual care; for patients randomized to usual care, we predicted the likelihood of AF diagnosis had they been randomized to screening. In each arm of the trial, we fit a generalized linear model (GLM) of the outcome (AF diagnosis) as a function of all the predictors described above using a Poisson distribution offset by the log of the follow-up time to account for differential follow-up time.^25^ This model was fit using the least absolute shrinkage and selection operator (LASSO), which we used to perform variable selection and parameter estimation via regularization.^26^ The LASSO hyperparameter (lambda) was determined using 10-fold cross-validation, and variable selection was entirely data-driven. We repeated this procedure twice, separately using the screening arm data only and again using the usual care data only. The model developed in the screening arm was used to predict AF likelihood under screening for participants randomized to usual care. The model developed in the usual care arm was used to predict AF likelihood under usual care for participants randomized to screening.

#### Step 2: Predict AF diagnosis for participants had they been randomized to their original arm using leave-one-out modeling

We used a set of models to predict AF diagnosis for participants given their actual randomization arm using leave-one-out cross-validation to limit bias.^27^ Starting with the usual care arm, we fit a model using all usual care participants except the *i*^th^ observation. As above, we used a GLM with Poisson distribution offset by the log of follow-up time, using LASSO for variable selection and parameter estimation via regularization. The hyperparameter from Step 1 was used. This model was used to predict the likelihood of AF diagnosis for the *i*^th^ observation in the usual care arm. This process was repeated 14774 times, once for each observation randomized to the usual care arm. The same procedure was repeated for observations randomized to the screening arm where 14882 iterative models were fit to predict the likelihood of AF diagnosis with screening.

#### Step 3: Calculate the individualized estimate of the effect of screening on AF diagnosis

These steps resulted in two predicted probabilities of AF diagnosis for each participant (n=29,656)—one had they been screened and one had they had usual care. The difference between these two was the individualized estimate of the effectiveness of screening. Crucially, both values were estimated using models that did not include the index observation.

### Risk-based approach to heterogeneity

In a second approach, we measured the heterogeneity of AF screening using a risk-based approach. We used the CHARGE-AF risk score to estimate the baseline risk of developing AF.^28^ CHARGE-AF is a risk model developed in the Atherosclerosis Risk in Communities study, Cardiovascular Health Study, and the Framingham Heart Study to predict incident AF. It uses age, race, height, weight, blood pressure, current smoking, use of antihypertensive medication, diabetes, myocardial infarction history, and heart failure as predictors. The CHARGE-AF model has been externally validated.^29^

### Testing for heterogeneity

We assessed heterogeneity by testing the interaction between the randomization arm (screening vs. usual care) and the predicted screening effectiveness (**Supplemental Methods 1**). Specifically, we fit a generalized linear model where the outcome of new AF diagnosis was a function of the randomization arm, the quartile of predicted screening effectiveness, and the interaction between the two. The model was fit using a Poisson distribution offset by the log of the follow-up time to account for differential follow-up time.^25^ We tested the statistical significance of the interaction using the likelihood ratio test. For visual representation we plotted the observed outcome rate by randomization arm, stratified by quartile of screening effectiveness.^14^ We repeated this procedure to test for heterogeneity by quartiles of predicted AF risk.

### Patient characteristics by predicted effectiveness and predicted risk

We used heatmaps to visualize participant characteristics by quartile of predicted screening effectiveness and predicted AF risk. We color-coded characteristics by their z-transformed value, where the darkest and lightest shade represent the highest and lowest value of a given patient characteristic, respectively.

### Correlation of predicted effectiveness and predicted risk

We plotted the percentile of predicted screening effectiveness against the percentile of predicted AF risk using hexagonal binning.^30^ To determine the correlation between the predicted effectiveness of screening and baseline risk, we calculated the Spearman correlation coefficient.

### Sensitivity Analyses

In a sensitivity analysis, we used a survival model (i.e., time-to-atrial fibrillation diagnosis) to test the robustness of the modified Poisson regression model used in the primary analysis. The analysis demonstrated no difference in time-to-atrial fibrillation diagnosis by randomization arm supporting the primary modeling approach (**Supplemental Methods 2**). Second, because CHARGE-AF was developed externally, its performance may be disadvantaged compared to the internally developed risk score. In a sensitivity analysis, we determined if an internally optimized AF risk model performed better than the CHARGE-AF model. We used the model to predict AF risk developed in the usual care arm (Step 1 above) as the internally optimized AF risk model.

## RESULTS

### Average effectiveness of screening

We present the baseline characteristics of 29,656 study participants in **Table 1**. Features were well balanced in the two arms of the trial. In 10,113 person-years of follow-up, 217 people in the usual care group were diagnosed with atrial fibrillation (2.15 per 100-person years). In 10,061 person-years of follow-up, 247 people in the screening group were diagnosed with atrial fibrillation (2.45 per 100 person-years). These rates were not statistically significantly different (rate difference 0.31 per 100 person-years, 95%CI −0.11 to 0.73).

**Table 1:** Patient characteristics by randomization arm.

| **Characteristic** | **Screening**<br>**(n=14882)** | **Usual care**<br>**(n=14774)** |
| --- | --- | --- |
| Age, mean (SD), yrs | 74 (7) | 74 (7) |
| Female sex, n (%) | 8,931 (60%) | 8,632 (58%) |
| Race, n (%) |  |  |
| White | 12,303 (83%) | 12,234 (83%) |
| Black | 771 (5.2%) | 681 (4.6%) |
| Hispanic | 324 (2.2%) | 295 (2.0%) |
| Other | 1,484 (10.0%) | 1,564 (11%) |
| Systolic blood pressure, mean (SD), mmHg | 131 (16) | 131 (17) |
| Diastolic blood pressure, mean (SD), mmHg | 75 (9) | 74 (9) |
| Heart rate, mean (SD), bpm | 75 (13) | 74 (13) |
| Weight, mean (SD), kg | 76 (18) | 76 (18) |
| Height, mean (SD), cm | 165 (10) | 165 (10) |
| English language preference, n (%) | 13,566 (91%) | 13,144 (89%) |
| Current smoking, n (%) | 709 (4.8%) | 768 (5.2%) |
| Hypertension, n (%) | 11,230 (75%) | 11,164 (76%) |
| Coronary artery disease, n (%) | 3,030 (20%) | 2,945 (20%) |
| Diabetes, n (%) | 3,510 (24%) | 3,464 (23%) |
| Congestive heart failure, n (%) | 1,424 (9.6%) | 1,400 (9.5%) |
| Prior stroke, n (%) | 1,201 (8.1%) | 1,204 (8.1%) |
| Vascular disease, n (%) | 2,835 (19%) | 2,801 (19%) |
| Anemia, n (%) | 3,599 (24%) | 3,640 (25%) |
| Bleeding history, n (%) | 5,467 (37%) | 5,739 (39%) |
| Chronic kidney disease, n (%) | 2,018 (14%) | 2,271 (15%) |
| Antihypertensive medication, n (%) | 10,357 (70%) | 10,402 (70%) |
| Oral anticoagulant use, n (%) | 324 (2.2%) | 339 (2.3%) |
| Rate control medication use, n (%) | 7,005 (47%) | 7,029 (48%) |
| Antiarrhythmic medication use, n (%) | 38 (0.3%) | 37 (0.3%) |
| 12 lead ECG in prior year, n (%) | 4,647 (31%) | 4,643 (31%) |
| PCP visits in the prior year, mean (SD) | 4.0 (4.5) | 4.6 (4.6) |
| CHARGE-AF score, mean (SD) | 13.45 (0.94) | 13.46 (0.95) |

### Model performance

The constituent models used to estimate the screening effectiveness discriminated moderately well (c-statistic 0.72 when predicting AF in usual care conditions; 0.73 when predicting AF in screening conditions) (**Figures S3**). In the usual care model, using the 75^th^ percentile as a threshold (diagnosis rate >1.87 per 100 person-years), the model specificity was 0.75 and the sensitivity was 0.55 (**Table S1**). In the screening model, using the 75^th^ percentile as a threshold (diagnosis rate >2.09 per 100 person-years), the model specificity was 0.76 and the sensitivity was 0.57. Visually, the model to predict AF diagnosis in the usual care arm was well calibrated and the model to predict AF diagnosis in the screening arm overpredicted AF risk at the low end of the predicted risk range (**Figure S4**).

The CHARGE-AF risk model also discriminated moderately well (c-statistic 0.73 in the usual care arm; 0.74 in the screening arm) (**Figure S5**). Using the CHARGE-AF model, using the 75^th^ percentile as a threshold, the predicted diagnosis rate in the usual care arm was >1.67 per 100 person-years, and >1.62 per 100 person-years in the screening arm. At this threshold, in both arms, the model specificity was 0.76 and the sensitivity was 0.60 (**Table S1**). Visually, the CHARGE-AF model substantially overpredicted AF risk at the highest risk range of the usual care arm but was well calibrated in the screening arm (**Figure S6**).

### Effect-based approach to screening heterogeneity

For participants whose predicted screening effectiveness was in the highest quartile, screening was associated with a statistically significant increase in AF diagnoses (4.00 vs. 2.88 per 100 person-years, rate difference 1.12 per 100 person-years, 95% CI 0.11 to 2.13) (**Figure 1**). In the remaining quartiles, the observed rates of AF diagnosis in the screening and usual care groups were not significantly different. There was a monotonic increase in observed screening effectiveness and interaction was statistically significant (p-value 0.01). The observed effectiveness of screening was concordant with the predicted effectiveness of screening in each quartile (**Figure S7**).

**Figure 1:**
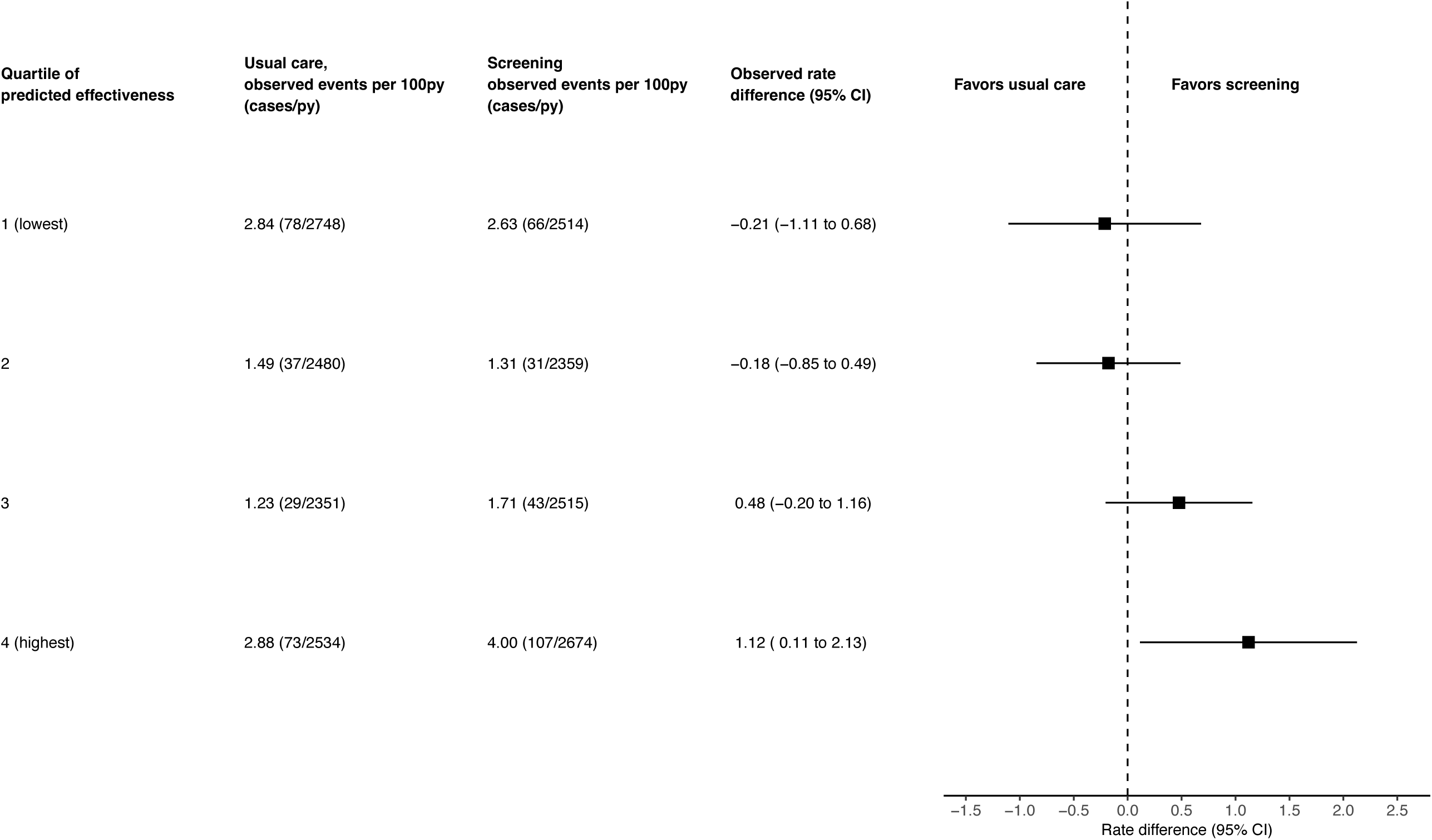
Heterogeneity of Screening Effectiveness using an Effect-based Model. <u>Legend</u> py–person-years. Model interaction p-value = 0.01

### Risk-based approach to screening heterogeneity

For participants whose predicted AF risk was in the highest risk quartile, screening was associated with a statistically significant increase in AF diagnoses (5.55 vs. 4.23 per 100 person-years, rate difference 1.32 per 100 person-years, 95% CI 0.14 to 2.5) (**Figure 2**). In the remaining quartiles, the observed rates of AF diagnosis in the screening and usual care groups were not significantly different. There was not a monotonic increase in observed screening effectiveness, and the interaction was not statistically significant (p-value 0.28). In a sensitivity analysis, we determined that an internally optimized risk model (i.e., the AF risk model from the usual care arm) did not identify a subgroup where screening was effective (**Figure S9**).

**Figure 2:**
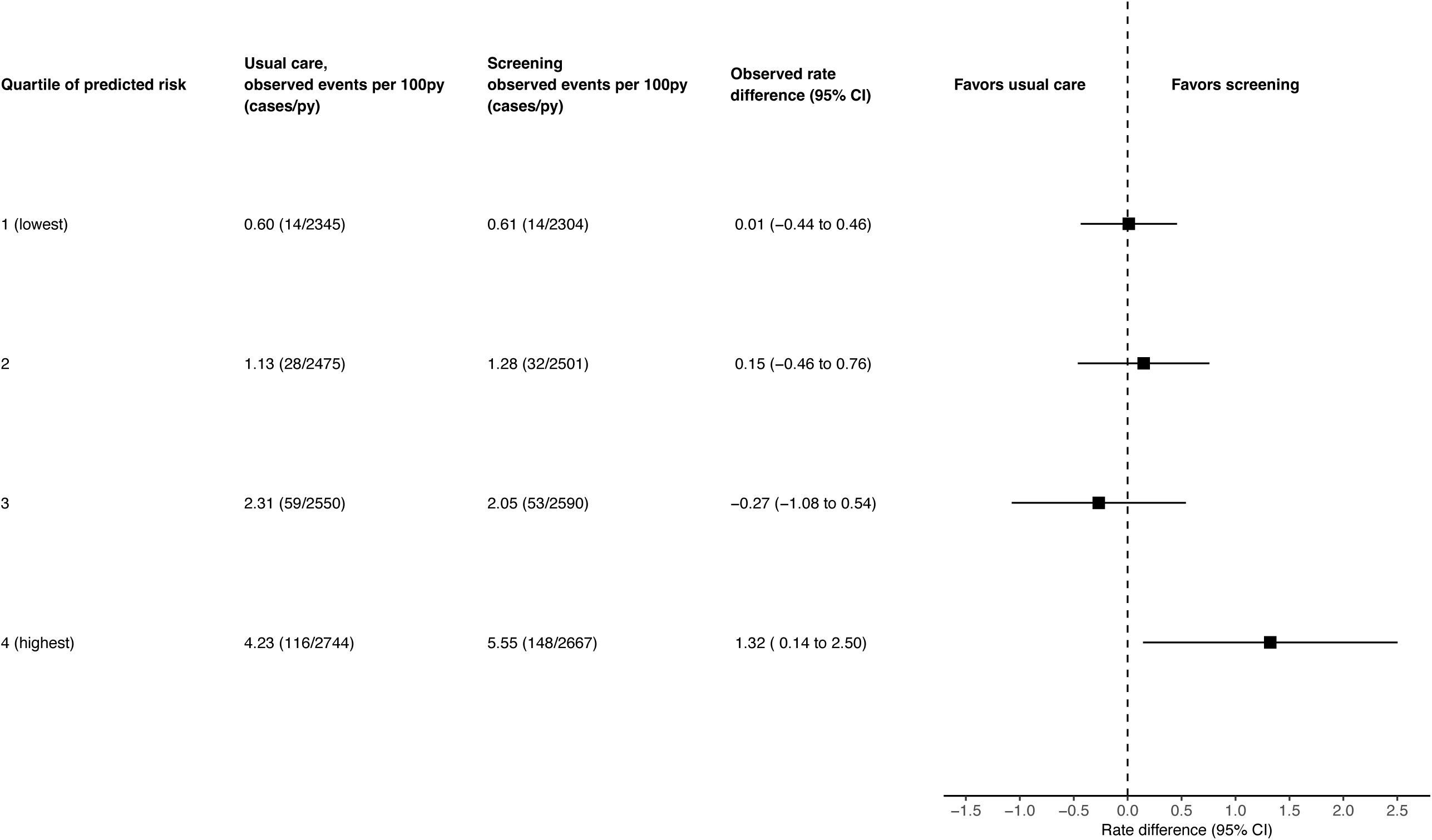
Heterogeneity of Screening Effectiveness using a Risk-based Model. <u>Legend</u> py–person-years. The baseline risk of incident AF was predicted using the CHARGE-AF score. Model interaction p-value = 0.28

### Patient characteristics of effective screening groups

In **Figure 3**, we display patient characteristics by quartile of screening effectiveness. Participants with higher BMI, greater number of PCP visits in the prior year, and higher heart rates were overrepresented in the lowest screening effectiveness quartile. Advanced age, high systolic blood pressure, and rate control medication use were more common in the highest quartile of screening effectiveness. Several characteristics displayed a U-shaped relationship, with high proportions in both the lowest and highest effectiveness quartile, such as hypertension, 12-lead ECG in the prior year, and congestive heart failure.

**Figure 3:**
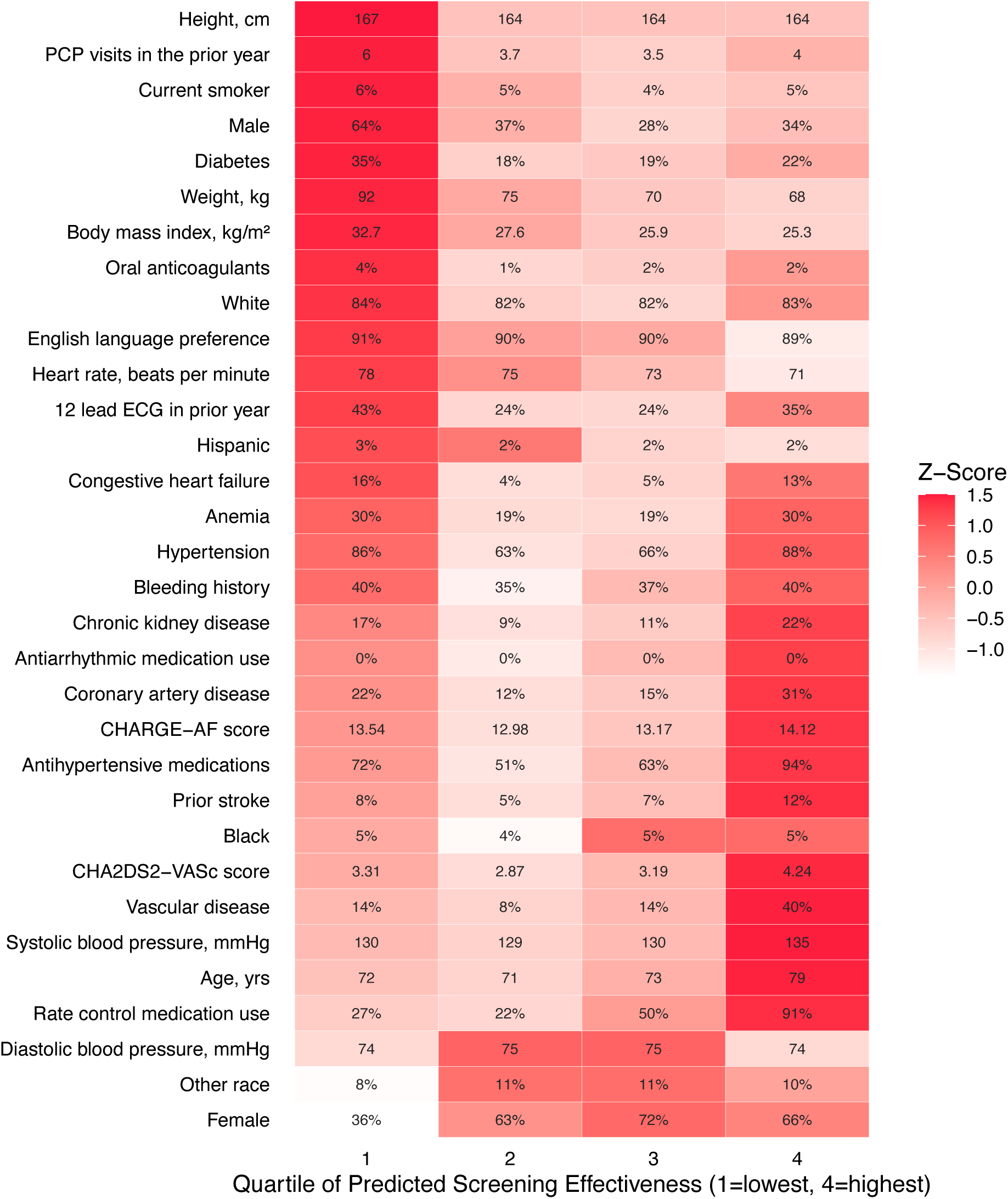
Heatmap of patient characteristics by quartile of predicted screening effectiveness. <u>Legend</u>: ECG – electrocardiogram, PCP – primary care physician, CHARGE-AF - Cohorts for Heart and Aging Research in Genomic Epidemiology atrial fibrillation. We present the average for continuous measures, and for categorical measures, we present the percent. We color-coded characteristics by their z-transformed value where the darkest and lightest shade represents the highest and lowest value of a given patient characteristic, respectively.

### Patient characteristics of risk groups

In **Figure 4**, we display the patient characteristics by decile of AF risk estimated using the CHARGE-AF score. Black participants, Hispanic participants, and women were overrepresented in the lowest-risk quartile. As expected, predictors used to calculate the CHARGE-AF score were more common in the highest risk quartile, including older age; higher height, weight, or blood pressure; smoking history; White racial identity; antihypertensive medication use; or diagnosis of diabetes, CHF, or prior MI. Among variables not directly used to calculate CHARGE-AF, we found that participants in the highest risk quartile were more likely to be male, have chronic kidney disease, and anemia.

**Figure 4:**
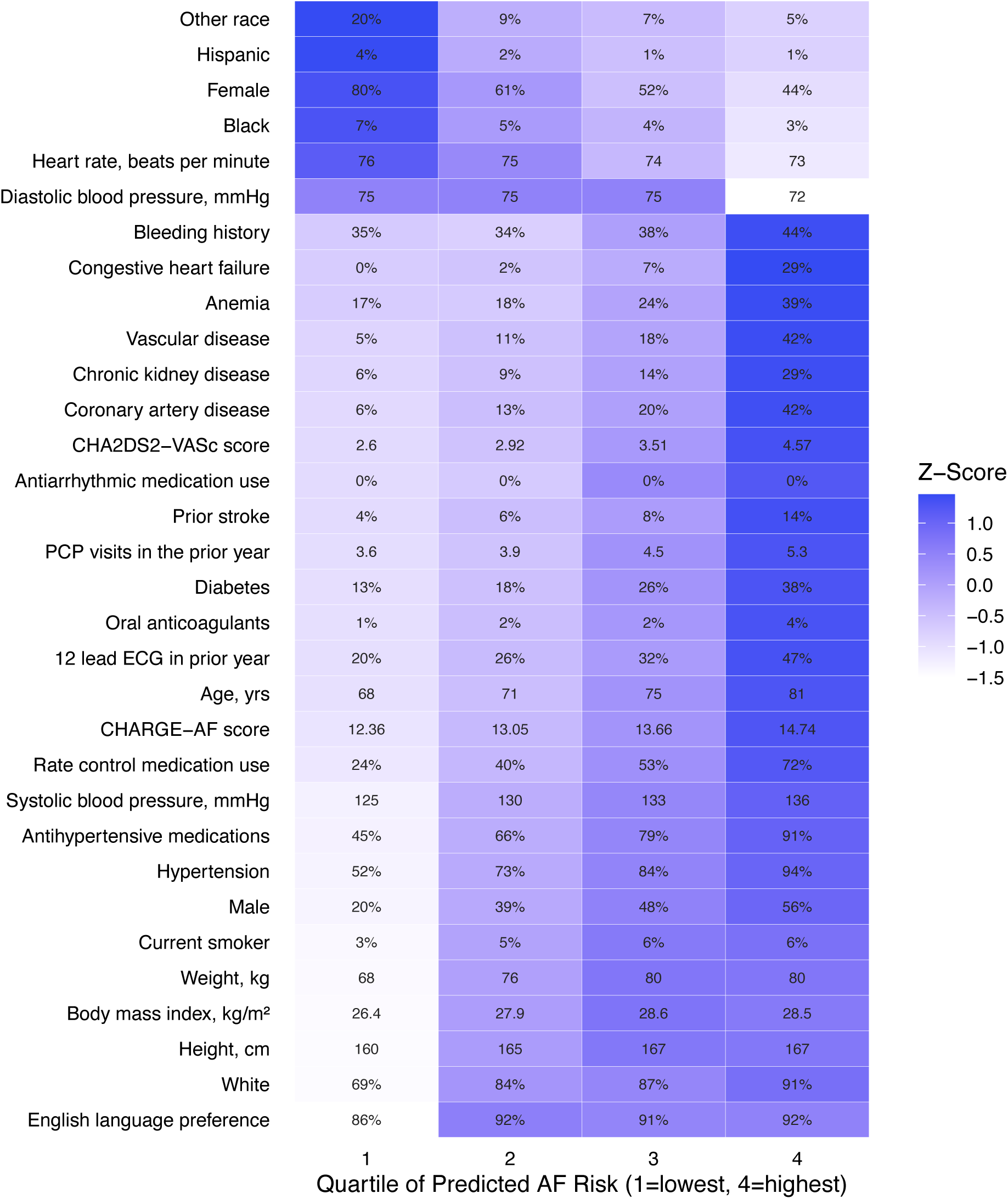
Heatmap of patient characteristics by quartile of CHARGE-AF score, predicted risk of AF. <u>Legend</u>: ECG – electrocardiogram, PCP – primary care physician, CHARGE-AF - Cohorts for Heart and Aging Research in Genomic Epidemiology atrial fibrillation. We present the average for continuous measures, and for categorical measures, we present the percent. We color-coded characteristics by their z-transformed value where the darkest and lightest shade represents the highest and lowest value of a given patient characteristic, respectively.

### Relationship between predicted risk and predicted screening effectiveness

The predicted risk of AF and the predicted AF screening effectiveness have a non-linear relationship. In **Figure 5**, we plot the relationship between the percentile of baseline AF risk measured by the CHARGE-AF score against predicted AF screening effectiveness. Predicted AF risk and predicted screening effectiveness were weakly correlated (Spearman correlation coefficient 0.23).

**Figure 5:**
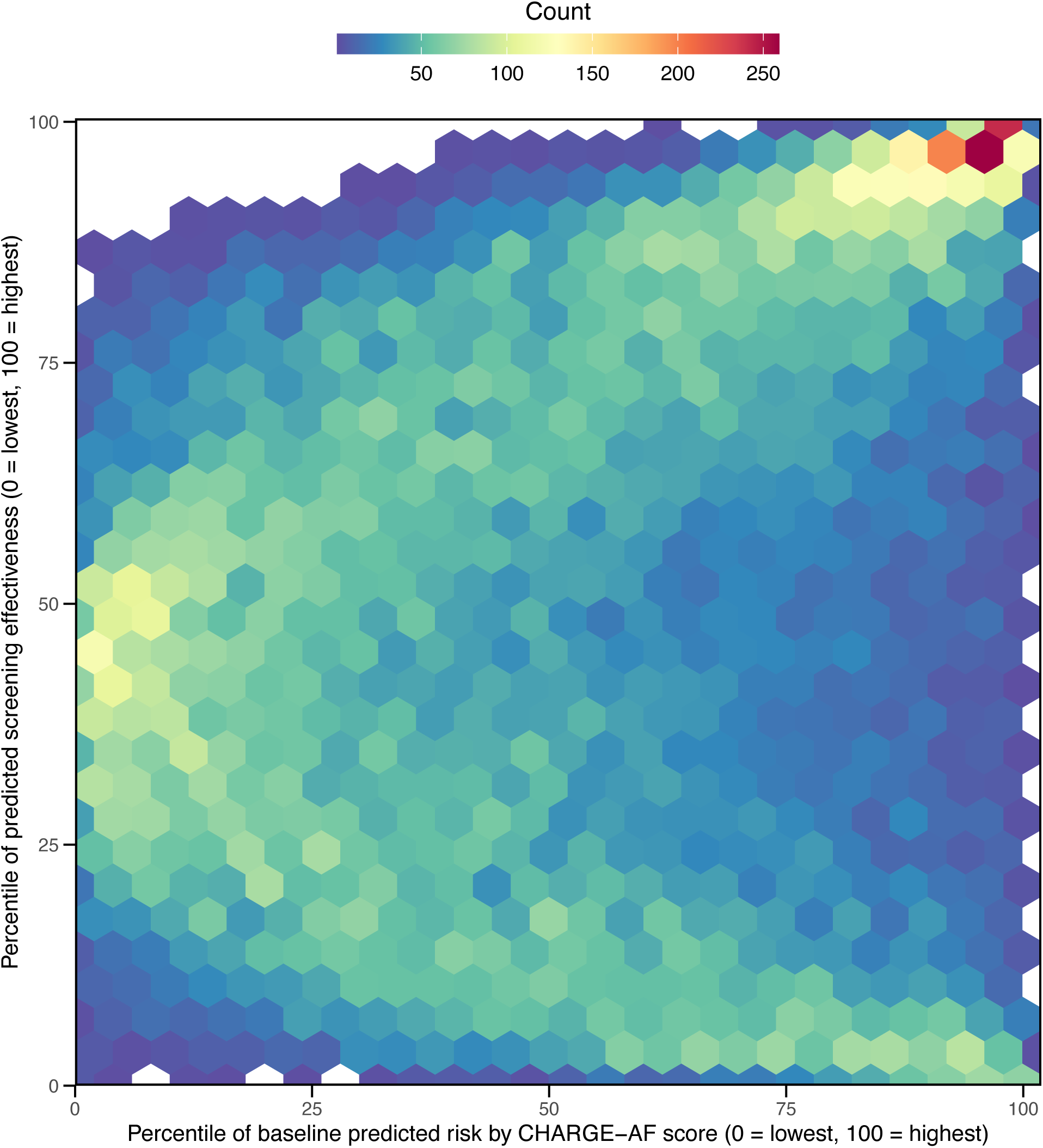
Correlation of predicted AF risk and predicted screening effectiveness. <u>Legend</u>: Each hexagonal tile represents study participants for a given percentile of baseline risk by CHARGE-AF score and the corresponding percentile of predicted screening effectiveness. The plot uses hexagonal binning to visualize the density of the bivariate data in a scatter plot.^30^ The color represents the density of individuals at that intersection (blue, green, yellow, orange, red indicating lowest density to highest density). The Spearman correlation coefficient is 0.23.

In the study, 11,050 (37%) were in either the high-effectiveness quartile or the high-risk quartile. In this group, 3,778 were in the high-effectiveness/high-risk group, while 3,636 were in the high-effectiveness/low-risk group, and 3,636 were in the low-effectiveness/high-risk group. Fully 18,606 (63%) patients were in the low-effectiveness/low-risk group (**Table S2**). Compared to the low-effectiveness/high-risk group, participants in the high-effectiveness/low-risk group were younger (mean age 75 vs. 79 years), had fewer visits in the prior year (mean visits 3.5 vs. 6.2), and were more likely to be female (77% vs. 32%), and Black (6.7% vs. 2.7%) (**Table S3**). Additionally, they were less likely to have hypertension (81% vs. 93%), diabetes (14% vs. 45%), or congestive heart failure (2.6% vs. 35%).

## DISCUSSION

In this secondary analysis of the VITAL-AF randomized trial, we identified subgroups in whom brief one-time screening was effective using both an effect-based modeling approach and a risk-based approach. Individuals identified through the effect-based approach were often females, in their late 70s, had hypertension, used rate control medications, and were seen less often by their PCP—a complete phenotype is available online.^31^ Individuals identified through the risk-based approach were often males, in their 80s, had hypertension, and were often seen by their PCPs. While there was overlap between high-risk and high-effectiveness groups, predicted risk and predicted screening effectiveness were only weakly correlated. This suggests that when screening for AF with a brief one-time screen, predicted risk is an incomplete surrogate for screening effectiveness.

There are a few possible reasons for the observed discordance between AF risk and screening effectiveness. First, the effectiveness model may identify people who do not fit physicians’ heuristic for AF, known as representation bias.^32,33^ For example, non-White racial identity, females, and those with a language preference other than English were over-represented in the high-effectiveness group. These same characteristics were more prevalent in low-risk groups. Second, screening effectiveness may be a function of healthcare access and connectedness.^34^ The average number of PCP visits in the prior year was lowest in the high-screening effectiveness group. This indicates that individuals who are often seen by their PCP may benefit less from one-time screening despite being at high risk for developing AF, presumably because physicians have an increased opportunity to detect heart rhythm abnormalities. By extension, one-time screening may be more effective for people with limited healthcare access. Third, it is possible that those in the high-effectiveness group are more likely to develop asymptomatic AF. For example, participants in the high-effectiveness group were considerably more likely to use medications that have rate control activity (e.g., beta blockers and calcium channel blockers). As a result, they may be more likely to have AF without rapid ventricular rate or marked palpitations. While this hypothesis is consistent with the study results, we could not test it with the available data.

The results of this study can inform future efforts at targeting screening in at least three ways. First, this study identified subgroups in which screening appears particularly effective. While VITAL-AF, D2AF, and Morgan & Mant found no average benefit of one-time screening, SAFE found an increase in AF diagnosis rate of 0.55% over 12 months.^6–8,21^ In our current study, we identified subgroups in whom screening was twice as effective as the average effect observed in SAFE. Targeting such subgroups may add efficiency to screening since most older individuals (63% in this study) are in the low-risk and low-effectiveness subgroup. Future trials targeting screening to the high-risk and high-effectiveness phenotype are needed to validate this strategy. There are shortcomings to this approach, namely that the subset in whom screening is effective is relatively small, and any targeting approach creates implementation challenges. To facilitate the identification of high-risk or high-effectiveness patients, we have published a supplemental online code that can be incorporated into electronic medical records or screening trials.^31^ Second, our finding that AF risk and screening effectiveness are weakly correlated indicates that screening trials targeting only high-risk individuals will not identify all individuals for whom screening is effective. Indeed, multiple trials are underway to test the value of screening by enrolling high-risk individuals.^35–38^ Third, our findings highlight the potential for inequity using a risk-based approach, particularly a risk equation like CHARGE-AF that uses race as a predictor. In this study, while non-White and female participants were concentrated in the low-risk quartiles, both groups were more evenly distributed across the spectrum of screening effectiveness. Thus, well-meaning efforts to target screening to high-risk individuals may widen disparities.

Finally, this study adds to the ongoing development of effect-based approaches to measuring the heterogeneous effect of cardiovascular interventions.^13–16^ For the treatment and prevention of many cardiovascular conditions, like statins to prevent atherosclerotic heart disease and anticoagulants to prevent stroke in AF, clinicians are asked to identify a subset of people for whom treatment is particularly effective.^39,40^ The conventional approach has been to model risk—that is, to develop an outcome model in untreated individuals and assume that this risk is tightly correlated with the effectiveness of the intervention, the true value of interest. This study demonstrates the vulnerability of this assumption. Prior studies have shown that risk-based models can effectively measure variation in treatment effect.^41,42^ This study showed that an effect-based approach can identify some individuals that a risk-based approach may miss.

Our study design and data source have important limitations. First, the high-effectiveness subgroup identified in this study needs to be externally validated. While we took caution when identifying the subgroup (i.e., using regularized regression models and leave-one-out cross-validation), external validation in a separate RCT is necessary to test the robustness of the findings. Second, this analysis was not pre-specified in the statistical analysis plan of the main trial. Thus, the results should be regarded as exploratory and hypothesis-generating. Third, screening for atrial fibrillation is important in so far as it prevents ischemic strokes. While one-time screening tends to identify individuals with high-burden AF, the full study was not powered to detect the effectiveness of screening on strokes and bleeding. Finally, AF screening spans multiple modalities (e.g., implantable cardiac monitors, smart watches) and venues (e.g., office-based, population-wide), each with its unique strengths and challenges.^43–46^ Even though most preventive screening occurs in primary care settings and single-lead screening was successfully performed in over 90%, the study findings may not generalize to other screening approaches.

Prior trials of brief, one-time AF screening have demonstrated mixed results. In a secondary analysis, we identified subsets where such screening was associated with increased AF diagnoses—one using a risk-based approach and another using an effect-based approach. While both approaches identified individuals for whom screening may be effective, predicted risk and predicted effectiveness were only weakly correlated, suggesting that AF risk is an incomplete proxy for the effectiveness of AF screening. Our effect-based approach identified that individuals with fewer office visits, perhaps reflecting barriers to care, and those using rate control medications that may mute symptoms of AF, benefited from screening even when they were not identified as high-risk for AF by standard risk models. Future AF screening efforts should focus on screening both “high-risk” and “high-effectiveness” people.

## Data Availability

Data are not publicly available.

## Non-standard Abbreviations and Acronyms

AF: atrial fibrillation
CHARGE-AF: Cohorts for Heart and Aging Research in Genomic Epidemiology AF
D2AF: Diagnosing Atrial Fibrillation
DAPT: Dual Antiplatelet Therapy
GLM: generalized linear model
LASSO: least absolute shrinkage and selection operator
PASCAL: PFO-Associated Stroke Causal Likelihood
PFO: patent foramen ovale
RCT: randomized clinical trial
SAFE: Screening for Atrial Fibrillation in the Elderly

## Author contributions

Dr. Shah had full access to all of the data in the study and takes responsibility for the integrity of the data and the accuracy of the data analysis. All authors listed have contributed sufficiently to the project to be included as authors, and all those who are qualified to be authors are listed in the author byline.

## SUPPLEMENTAL MATERIAL

**Figure S1:**
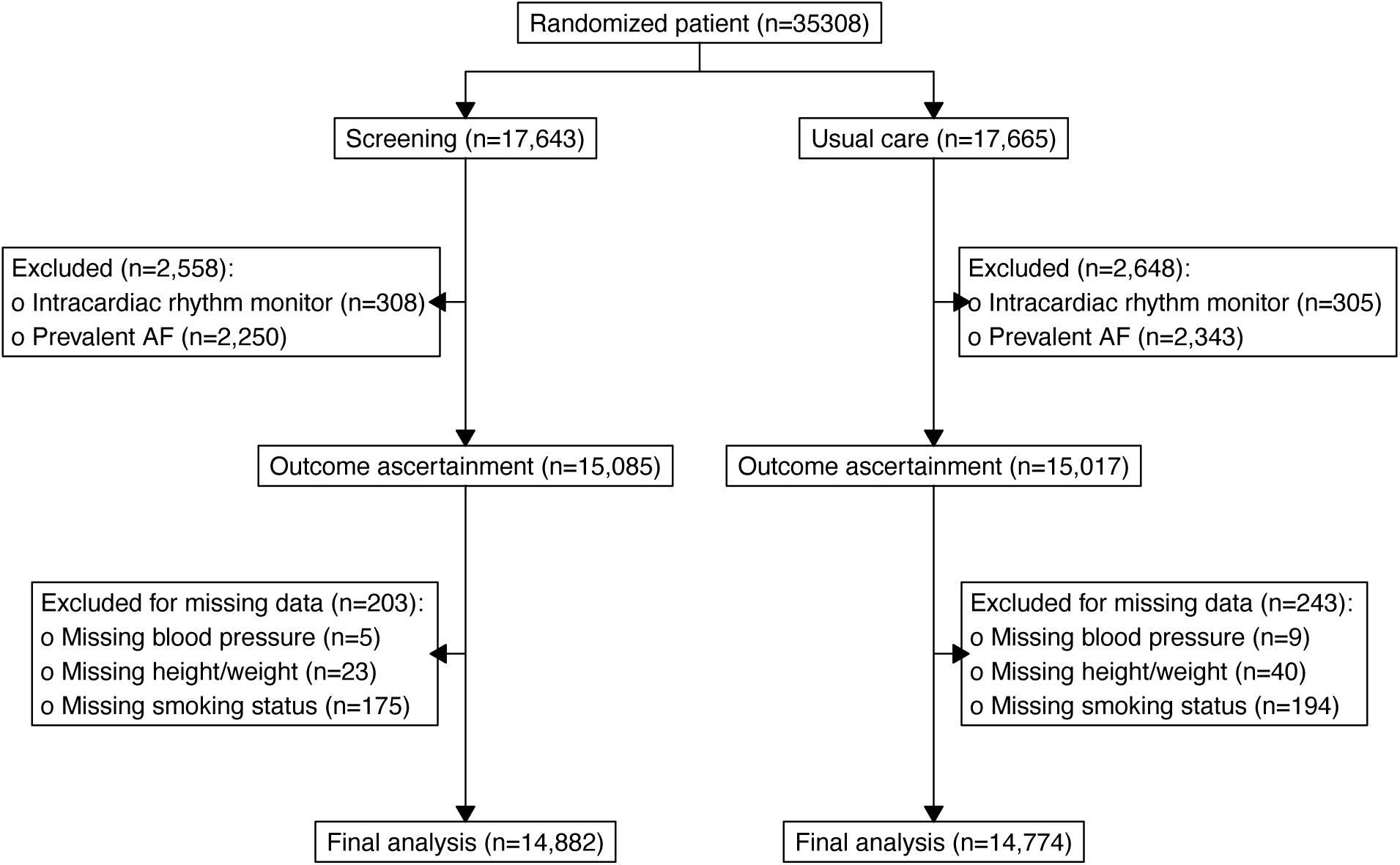
Consort diagram.

**Figure S2:**
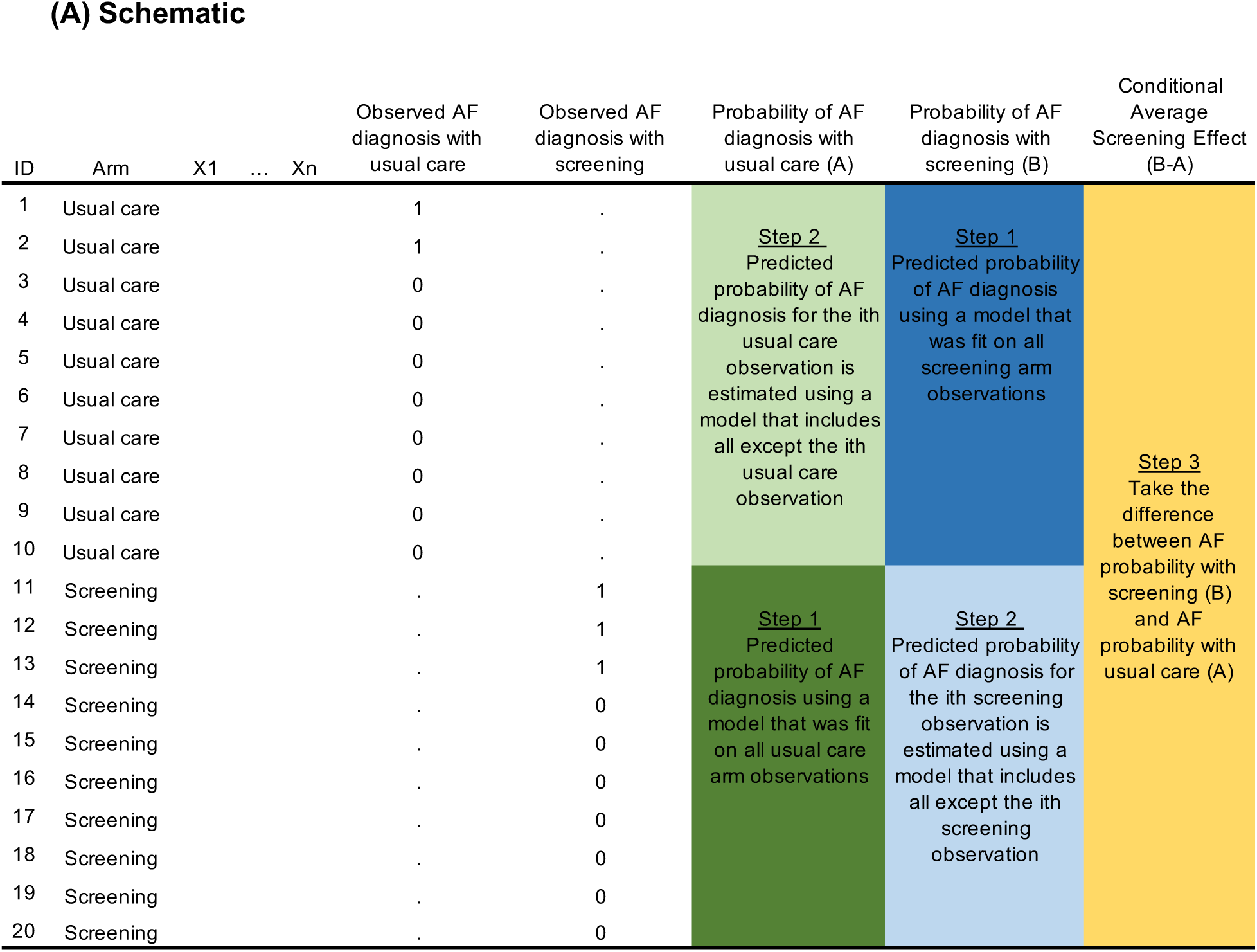
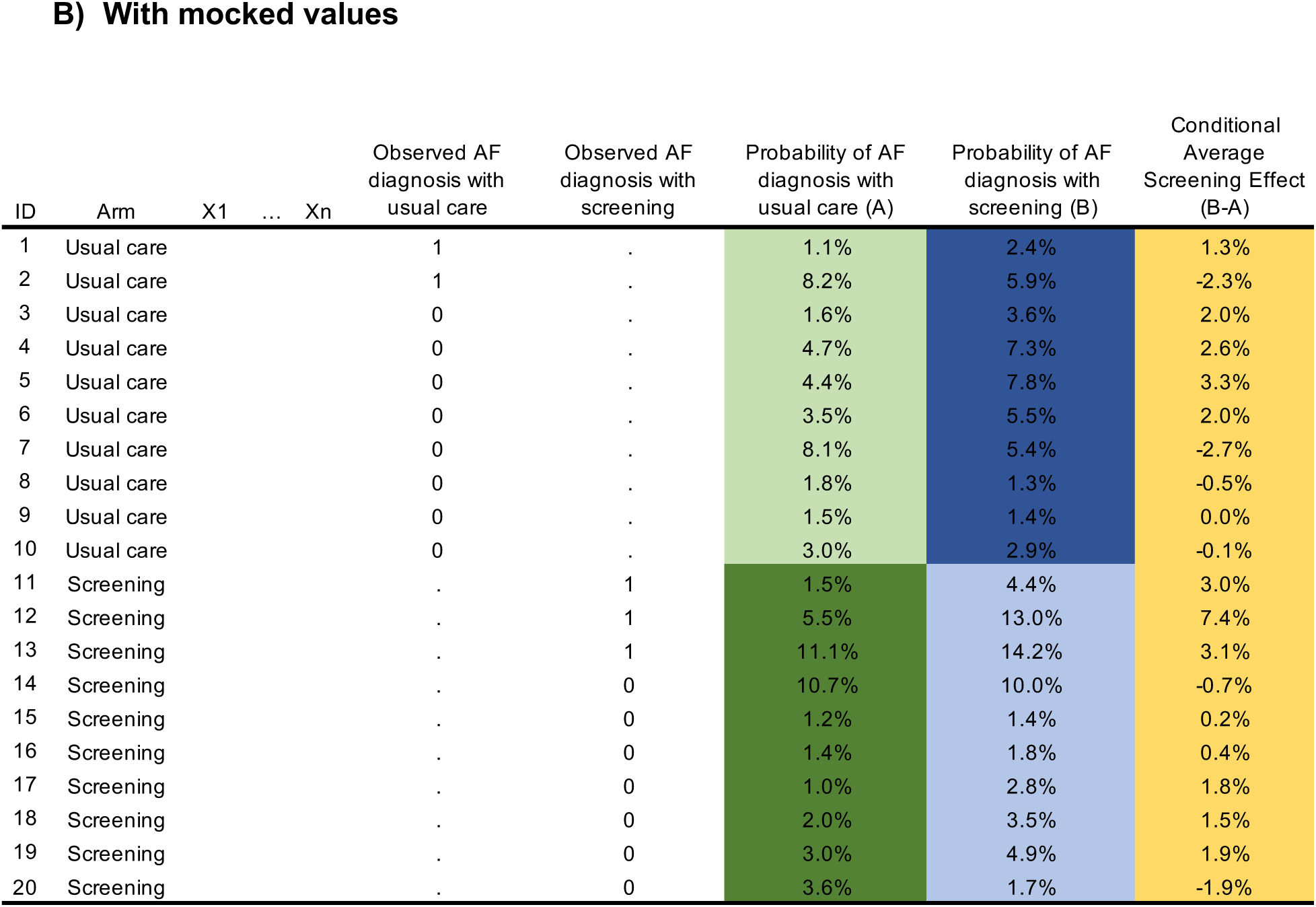
Visualization of modeling approach used to calculate the conditional average screening effectiveness. <u>Legend</u>: The two panels provide a visual representation of the modeling approach taken to calculate the conditional average screening effectiveness. The hypothetical data frame displays 20 observations, 10 randomized to usual care and 10 to screening. X1 to Xn represent predictors (not displayed). Outcomes are observed for each observation. Note that the outcome (AF diagnosis) under usual care can only be observed for those randomized to usual care, and the outcome under screening can only be observed for those randomized to screening. Through the methods described in the manuscript, we estimate two outcome probabilities for each observation, one had they been randomized to usual care, and the other had they been randomized to screening. The difference between these two is the conditional average screening effectiveness.

### Supplemental Methods 1 – Interaction testing

We tested for heterogeneity using interaction testing. The model used a Poisson distribution, and log-link with an offset for follow-up time. First, we fit a model with no interaction term:

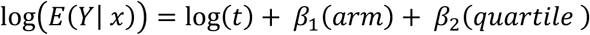

Where *t* is follow up time, *arm* takes values 0 for usual care and 1 for screening, and *quartile* takes values 1, 2, 3, or 4 based on the corresponding quartile of predicted screening effectiveness in the effect-based analysis and quartile of predicted risk in the risk-based analysis.

Second, we fit the following model with the interaction term:

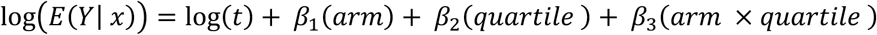

We compared the improvement in model fit when using an interaction term using the likelihood ratio test. The manuscript and figures report the p value from the likelihood ratio test.

### Supplemental Methods 2

We used a survival model with time-to-atrial fibrillation as the outcome to test the robustness of the modified Poisson regression model used in the primary analysis. The analysis demonstrated a linear rate of diagnosis and no difference in time-to-atrial fibrillation diagnosis by randomization arm supporting the primary modeling approach. The Kaplan-Meier curve demonstrates no statistical or clinically meaningful difference in time-to-AF diagnosis.

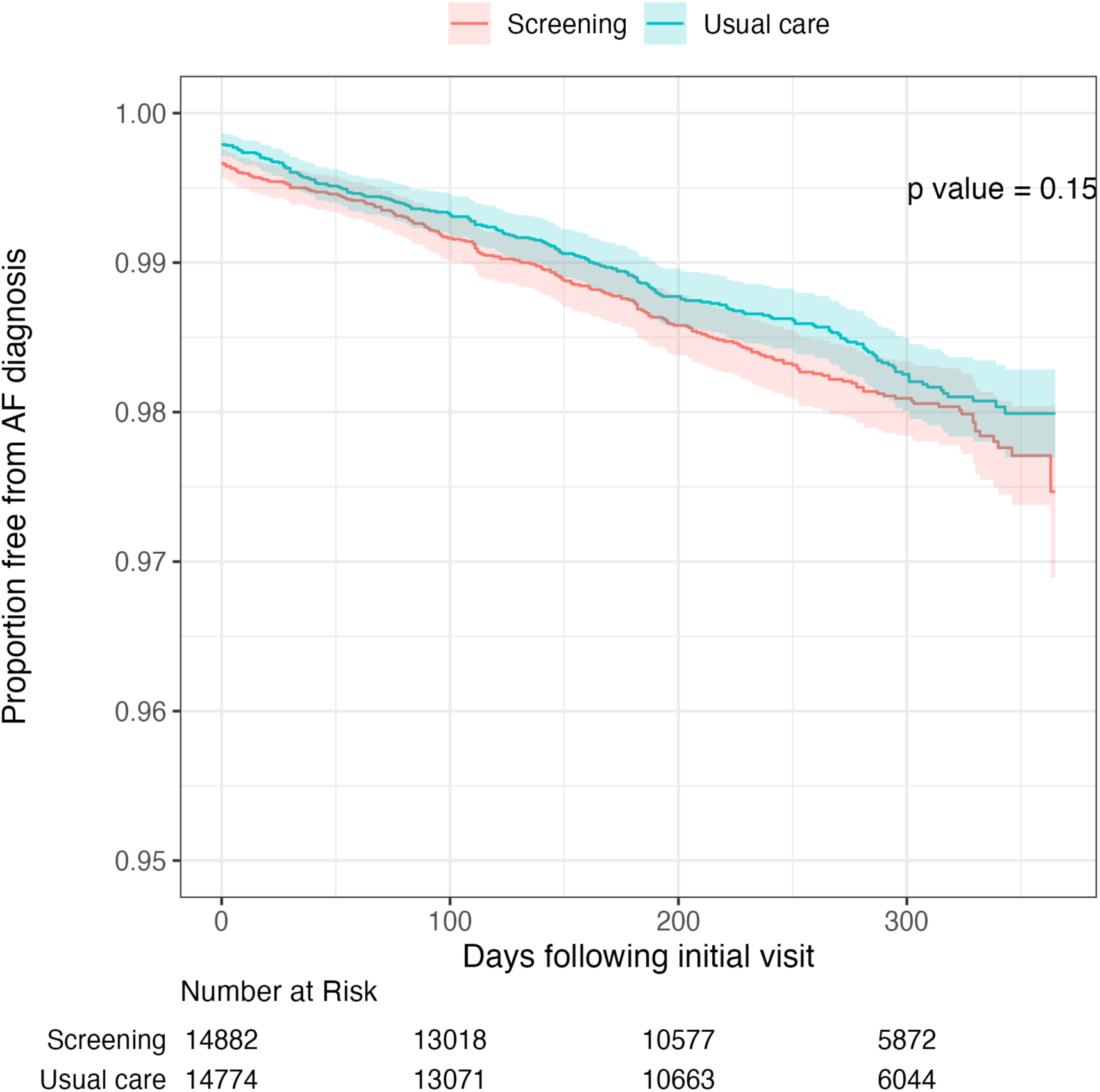

Further, among those with a new diagnosis of AF during the study follow-up, there was no statistical or clinically meaningful difference in the distribution of time to AF diagnosis.

|  | Screening<br>n = 262 | Usual care<br>n = 238 | p-value |
| --- | --- | --- | --- |
| Time to AF diagnosis, median (Q1, Q3) | 110 (13, 194) | 120 (30, 193) | 0.33 |

**Table S1:** Model performance at the 75^th^ percentile threshold.

| **Model approach** | **Arm** | **Classification threshold of AF diagnosis rate at 75<sup>th</sup> percentile** | **Specificity** | **Sensitivity** |
| --- | --- | --- | --- | --- |
| Components models used to calculate the of effectivenessscore (i.e., conditional average treatment effect) | Model to predict AF under usual care conditions (evaluated in those randomized to usual care) | >1.87 per 100 py | 0.75 | 0.55 |
|  | Model to predict AF under usual screening conditions (evaluated in those randomized to screening) | >2.09 per 100 py | 0.76 | 0.57 |
| CHARGE-AF model | Model to predict AF (evaluated in those randomized to usual care) | >1.67 per 100 py | 0.76 | 0.60 |
|  | Model to predict AF (evaluated in those randomized to screening) | >1.62 per 100 py | 0.76 | 0.60 |

**Figure S3:**
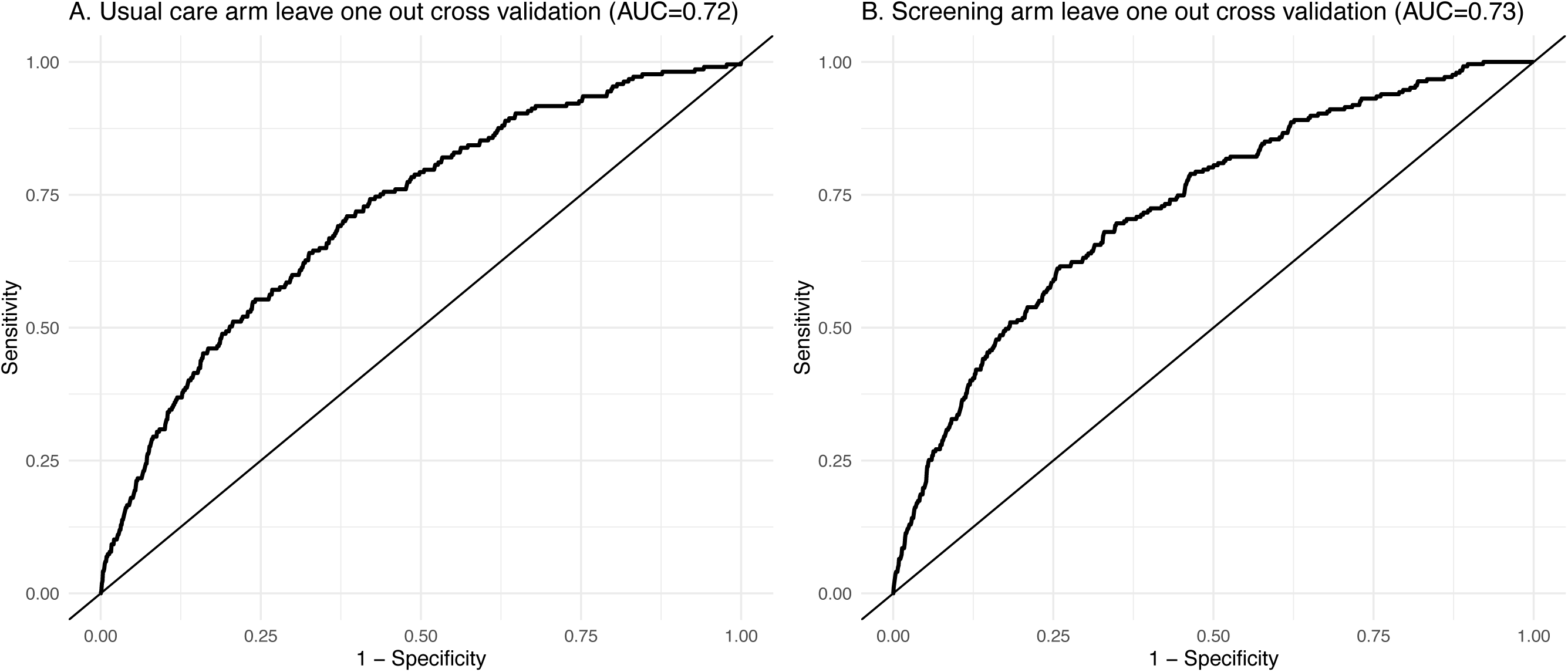
Receiver operator curves of constituent models used to calculate effectiveness scores using leave one out cross-validation.

**Figure S4:**
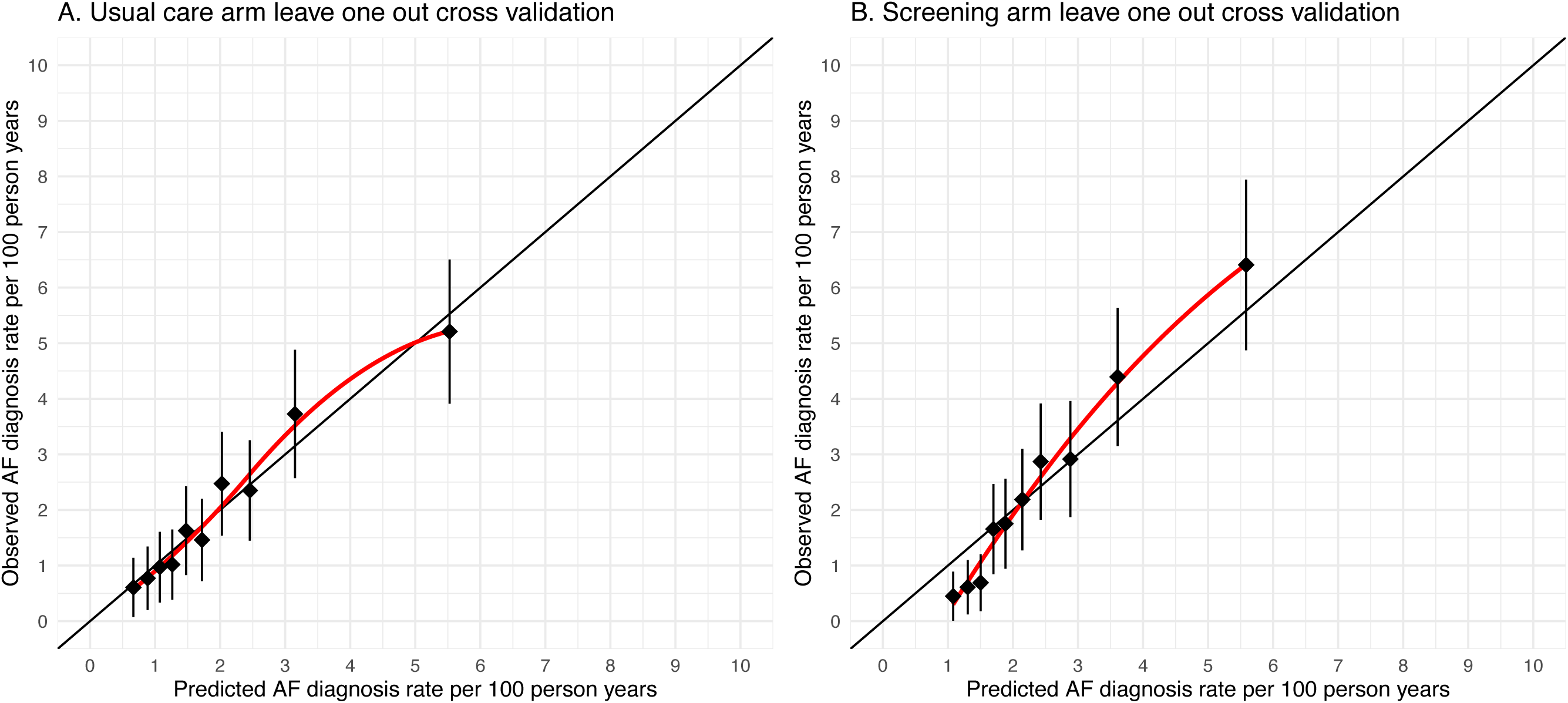
Calibration of constituent models used to calculate effectiveness scores using leave-one-out cross-validation.

**Figure S5:**
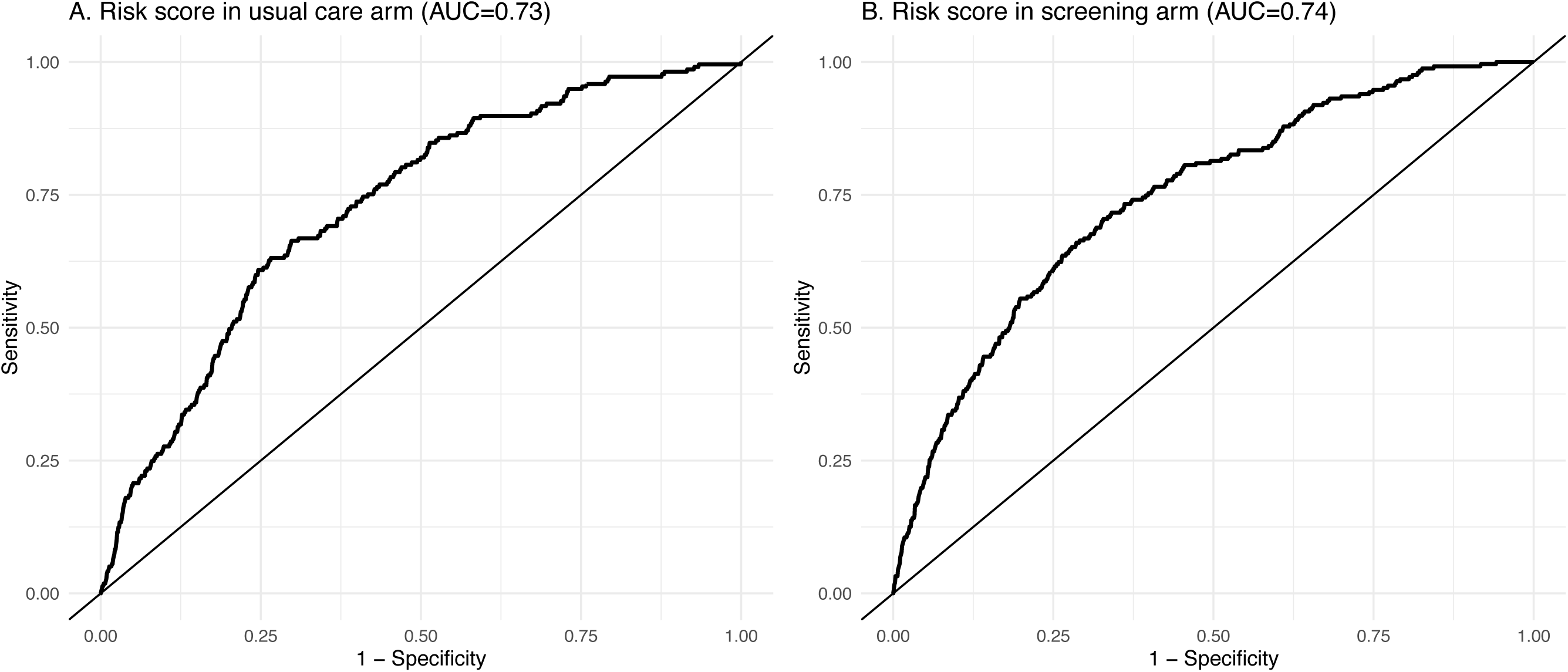
Receiver operator curves of CHARGE-AF risk model in usual care and screening arm.

**Figure S6:**
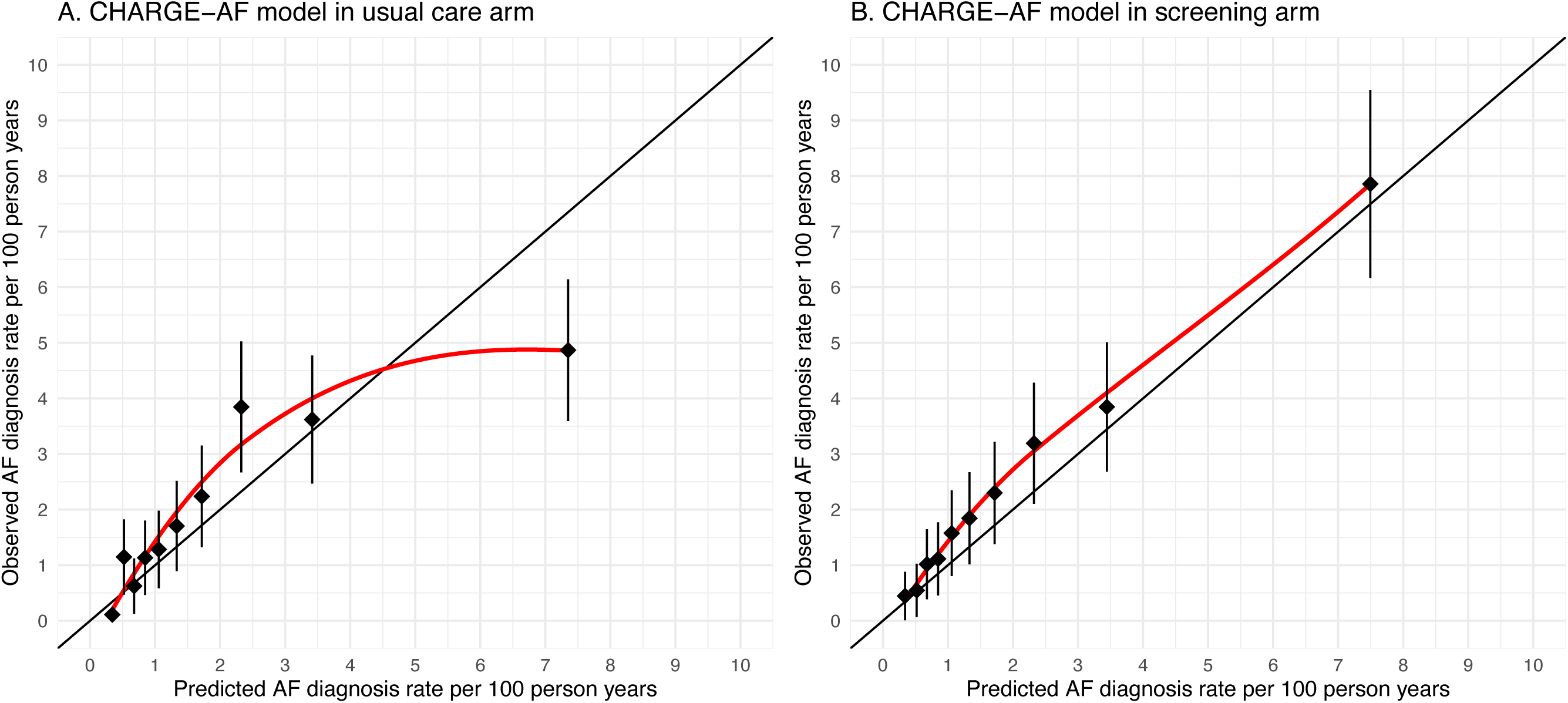
Calibration of CHARGE-AF risk model in usual care and screening arm.

**Figure S7:**
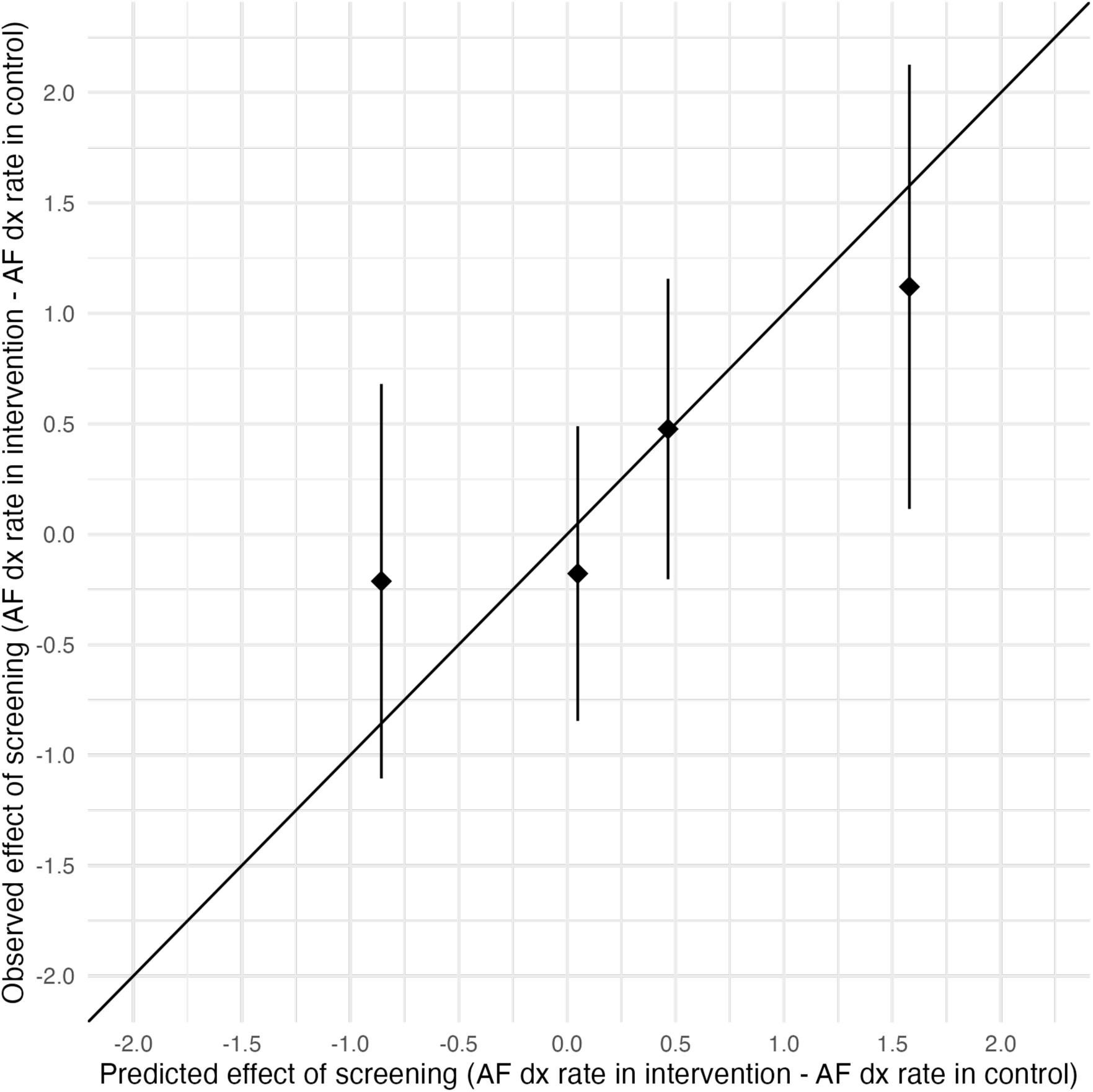
Observed vs. Expected Effectiveness of Screening by Quartile of Effect-based Score.

**Figure S8:**
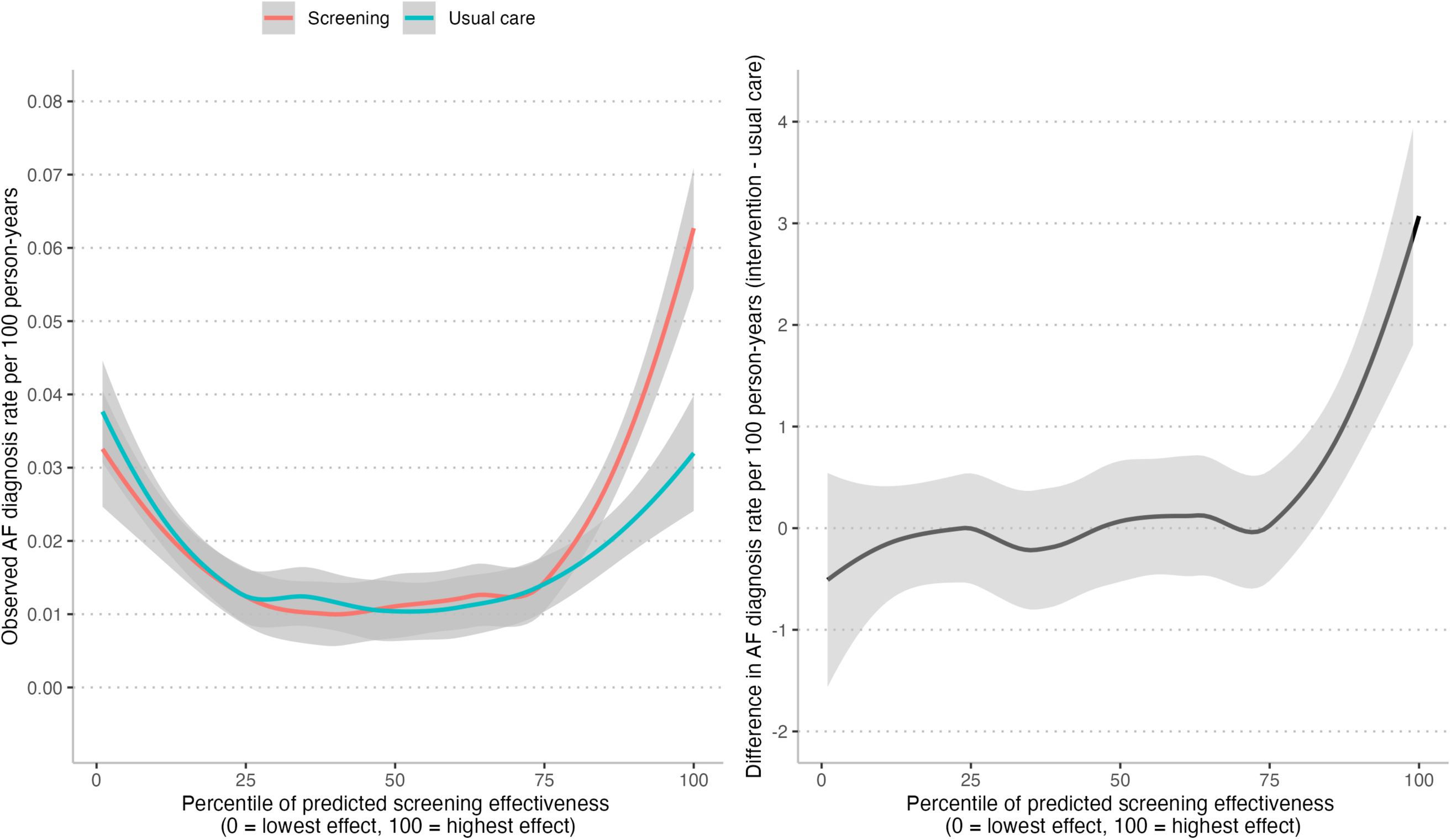
A Continuous Evaluation of the Effect-Based Model. <u>Legend</u>: This is a continuous representation of the data presented in Figure 1 (where it is presented as quartiles of predicted screening effectiveness). The left panel displays the diagnosis rate by randomization arm by percentile of predicted screening effectiveness. The line is a locally estimated best fit line (i.e., LOESS) with a span of 0.75. The right panel displays the difference in AF diagnosis rates; values above 0 indicate percentiles where screening is more effective than usual care.

**Figure S9:**
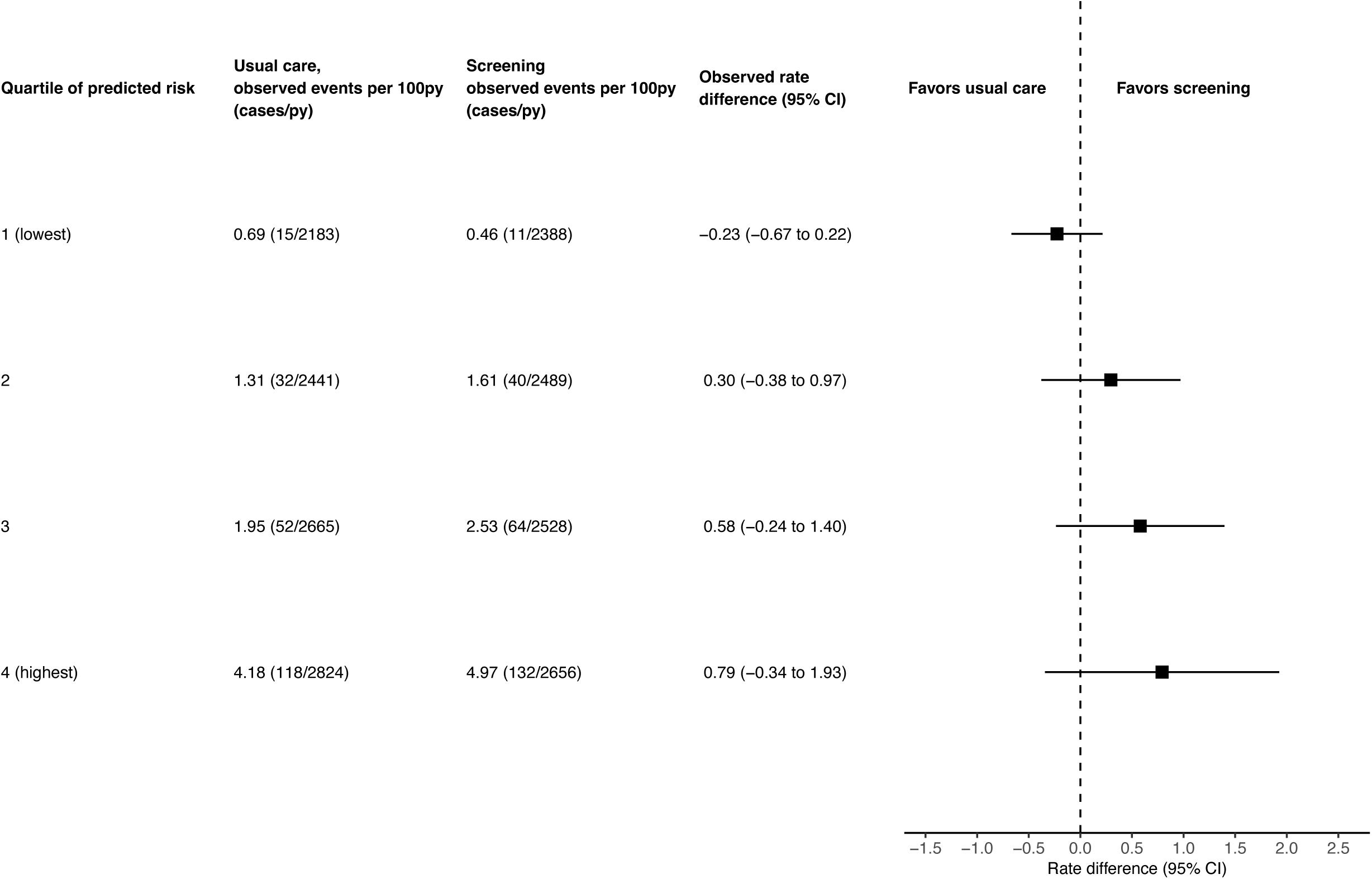
Heterogeneity of Screening Effectiveness using an Internally Developed Risk-based Model (i.e., AF risk model from the usual care arm) <u>Legend</u>: The CHARGE-AF score has a potential disadvantage because it was developed externally. We tested the robustness of our primary finding by using an internally optimized risk model. Recall that we created a risk model to estimate the likelihood of AF under usual care when estimating the effect-base score. We found that this usual care risk model and the CHARGE-AF score were highly correlated (Spearman correlation coefficient 0.84). Further, when we repeat the analysis, we find no heterogeneity of screening effectiveness when using the internally developed risk score (model interaction p =0.47).

**Table S2:** Contingency table of High Effectiveness and High Risk Quartiles.

|  | **High effectiveness quartile**<br>**(Q4)** | **Low effectiveness quartiles**<br>**(Q1-3)** | **Total** |
| --- | --- | --- | --- |
| **High risk quartile (Q4)** | 3778 (13%) | 3636 (12%) | 7414 |
| **Low risk quartiles (Q1-3)** | 3636 (12%) | 18606 (63%) | 22242 |
| **Total** | 7414 | 22242 | 29656 |

**Table S3:** Patient characteristics of those identified as high effectiveness but low risk compared to those identified as low effectiveness but high risk.

| **Characteristic** | **High effectiveness**<br>**&**<br>**Low risk**<br>**(n=3636)** | **Low effectiveness**<br>**&**<br>**High risk**<br>**(n=3636)** |
| --- | --- | --- |
| Age, mean (SD), yrs | 75 (5) | 79 (6) |
| Female sex, n (%) | 2,803 (77%) | 1,171 (32%) |
| Race, n (%) |  |  |
| White | 2,764 (76%) | 3,342 (92%) |
| Black | 242 (6.7%) | 98 (2.7%) |
| Hispanic | 81 (2.2%) | 39 (1.1%) |
| Other | 549 (15%) | 157 (4.3%) |
| Systolic blood pressure, mean (SD), mmHg | 132 (17) | 134 (17) |
| Diastolic blood pressure, mean (SD), mmHg | 75 (9) | 72 (9) |
| Heart rate, mean (SD), bpm | 72 (12) | 75 (13) |
| Weight, mean (SD), kg | 66 (12) | 89 (19) |
| Height, mean (SD), cm | 163 (9) | 169 (11) |
| English language preference, n (%) | 3,160 (87%) | 3,357 (92%) |
| Current smoking, n (%) | 138 (3.8%) | 257 (7.1%) |
| Hypertension, n (%) | 2,949 (81%) | 3,376 (93%) |
| Coronary artery disease, n (%) | 675 (19%) | 1,469 (40%) |
| Diabetes, n (%) | 505 (14%) | 1,644 (45%) |
| Congestive heart failure, n (%) | 95 (2.6%) | 1,260 (35%) |
| Prior stroke, n (%) | 315 (8.7%) | 464 (13%) |
| Vascular disease, n (%) | 1,037 (29%) | 1,182 (33%) |
| Anemia, n (%) | 759 (21%) | 1,390 (38%) |
| Bleeding history, n (%) | 1,300 (36%) | 1,591 (44%) |
| Chronic kidney disease, n (%) | 488 (13%) | 996 (27%) |
| Antihypertensive medication, n (%) | 3,400 (94%) | 3,120 (86%) |
| Oral anticoagulant use, n (%) | 57 (1.6%) | 161 (4.4%) |
| Rate control medication use, n (%) | 3,323 (91%) | 1,962 (54%) |
| Antiarrhythmic medication use, n (%) | 16 (0.4%) | 15 (0.4%) |
| 12 lead ECG in prior year, n (%) | 978 (27%) | 1,837 (51%) |
| PCP visits in the prior year, mean (SD) | 3.5 (3.1) | 6.2 (6.6) |
| CHARGE-AF score, mean (SD) | 13.37 (0.46) | 14.63 (0.53) |

